# Reliability of a smartphone app to objectively monitor performance outcomes in degenerative cervical myelopathy: an observational study

**DOI:** 10.1101/2024.02.10.24302007

**Authors:** Alvaro Yanez Touzet, Tatiana Houhou, Zerina Rahic, Angelos Kolias, Stefan Yordanov, David B. Anderson, Ilya Laufer, Maggie Li, Gordan Grahovac, Mark R. N. Kotter, Benjamin M. Davies, MoveMed AYT is the first

## Abstract

**Objectives:** To assess the reliability of the MoveMed battery of performance outcome measures.

**Design:** Prospective observational study.

**Setting:** Decentralised secondary care in England, United Kingdom.

**Participants:** Seven adults aged 59.5 (SD 12.4) who live with degenerative cervical myelopathy (DCM) and possess an approved smartphone.

**Primary and secondary outcome measures:** The primary outcome was to determine the test-retest reliability of the MoveMed performance outcomes using the intra-class correlation of agreement (ICC_agreement_). The secondary outcome was to determine the measurement error of the MoveMed performance outcomes using both the standard error of agreement of the mean (SEM_agreement_) and the smallest detectable change of agreement (SDC_agreement_). Criteria from the Consensus-based Standards for the selection of health Measurement Instruments (COSMIN) manual were used to determine adequate reliability (i.e., ICC_agreement_ ≥ 0.7) and risk of bias. Disease stability was controlled for using two minimum clinically important difference (MCID) thresholds obtained from the literature on the patient-derived modified Japanese Orthopaedic Association (P-mJOA): namely, MCID ≤1 point and MCID ≤2 points.

**Results:** All tests demonstrated moderate-to-excellent test-retest coefficients and low measurement errors. In the MCID ≤1 group, ICC_agreement_ values ranged from: 0.84–0.94 in the Fast Tap Test, 0.89–0.95 in the Hold Test, 0.95 in the Typing Test, and 0.98 in the Stand and Walk Test. SEM_agreement_ values ranged from ±1 tap, ±1–3% stability score points, ±0.06 keys per second, and ±10 steps per minute, respectively. SDC_agreement_ values were ±3 taps, ±4–7% stability score points, ±0.2 keys per second, and ±27 steps per minute. In the MCID ≤2 group, ICC_agreement_ values ranged from: 0.61–0.91, 0.75–0.77, 0.98, and 0.62, respectively; SEM_agreement_ values from: ±1 tap, ±2–4% stability score points, ±0.06 keys per second, and ±10 steps per minute; and SDC_agreement_ values from: ±3–7 taps, ±7–10% stability score points, ±0.2 keys per second, and ±27 steps per minute. Furthermore, the Fast Tap, Hold, and Typing Tests obtained sufficient ratings (ICC_agreement_ ≥ 0.7) in both MCID≤1 and MCID≤2 groups. No risk of bias factors from the COSMIN Risk of Bias checklist were recorded. Overall, ‘very good’ quality evidence of sufficient reliability was found for the MoveMed battery in DCM.

**Conclusions:** Criteria from COSMIN provide ‘very good’ quality evidence of the reliability of the MoveMed tests in an adult population living with DCM.

**Strengths and limitations of this study:**

- Criteria from the COSMIN manual support the reliability of MoveMed’s battery of performance outcome measures.
- Criteria from the COSMIN manual rate the methodological quality of the evidence at ‘very good’.
- Self-reported outcome measures and data elements were used in a decentralised secondary setting.
- Primary and secondary outcomes were used to assess reliability.
- Study design and analyses were performed by individuals formally trained in clinimetrics.

## INTRODUCTION

Degenerative cervical myelopathy (DCM) is a slow-motion spinal cord injury that is estimated to affect 1 in 50 adults (1, 2). In this disabling condition, dexterity, gait, and balance are key measurement constructs. As with numerous other diseases that affect the nervous and musculoskeletal systems, the monitoring of DCM today still largely depends on qualitative approaches such as hierarchical classifications of exemplar functions. While such measures can be robust, they remain intrinsically subjective. Hence, such measures pose significant challenges, especially in fluctuating diseases, as they are often unable to effectively detect small but significant changes in a timely fashion.

The current gold-standard measure in DCM is the modified Japanese Orthopaedic Association (mJOA) score but, unfortunately, it is found to possess low and inadequate responsiveness to detect disease change (3, 4). Variation, driven by both disease heterogeneity and instrument reliability, is more than twice the minimal clinically important difference (MCID). In practice, this demands sample sizes greater than 300 patients for 1:1 comparison with at least 80% power. Developing new approaches to functional measurement is, thus, a recognised research priority in DCM. Quantitative, quick, and decentralised disease monitoring could significantly improve control of heterogeneity and critical sample sizes; for instance, through use of performance-based outcome measures (PerfO’s or PerfOM’s).

MoveMed, a battery of PerfOM’s in a mobile application, recently demonstrated validity in DCM (5). The application leverages the accuracy of mobile sensors to assess hand, arm, and leg function in real-time, in the user’s natural environment, and under standardised conditions. It was designed by healthcare professionals from the University of Cambridge and is being developed in accordance with ISO 13485 (Software as a Medical Device). Its reliability, however, has not yet been formally assessed. Thus, in this study, we use formal methods and criteria from the United States Food and Drug Administration (FDA) and Consensus-based Standards for the selection of health Measurement Instruments (COSMIN) to assess the reliability of MoveMed’s PerFOM’s. This is the second article in a series of clinimetric studies on the measurement properties of MoveMed’s PerfOM’s (5).

## METHODS AND ANALYSIS

### Participants

Between September 2022 and April 2023, people living with a neurological condition were enrolled in the prospective and decentralised EMPOWER study (www.movemed.io/empower). Prospective participants were recruited via an online campaign and asked to complete consent and registration forms (Figure 1) (6, 7). These were used to screen participants for eligibility. Participants were deemed eligible if they had a self-reported diagnosis of DCM, owned a smartphone, and were able to stand and walk without the assistance of another person. Eligible participants were invited to download the MoveMed App to their smartphones and complete an electronic, baseline questionnaire on neuromuscular function, hand dominance, and quality of life. This included questions from the patient-derived mJOA (P-mJOA) and the World Health Organization Quality of Life Brief Version (WHOQOL-Bref).

**Figure 1.**
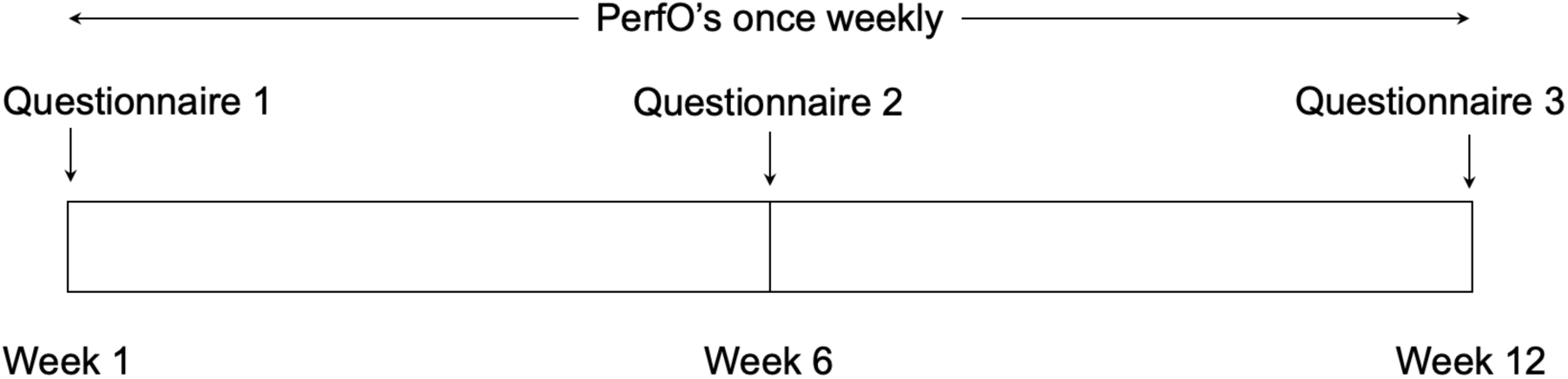
Study timeline.

All enrolled participants were asked to perform each task in the MoveMed App once per week for a period of 12 weeks. Task adherence was remotely monitored once a week using a bespoke online dashboard. Participants were offered reminders and help via email once a week if 14 days passed since the completion of the latest task. These were offered for a total of two consecutive times per participant, after which the participant was considered lost to follow-up. At weeks six and 12, participants were asked to complete the same electronic questionnaire from week one.

### MoveMed and tasks

The MoveMed App is a smartphone application designed by academic neurosurgeons and computer scientists from the University of Cambridge to administer PerfOM’s (Figure 2). These may be administered by clinicians during in-person visits or self-performed by individuals in community. Version 1.0.0 of the App originally offered three performance tasks: a Fast Tap Test, a Hold Test, and a Stand and Walk Test. Version 1.2.2 incorporated an additional offering—a Typing Test—while making no changes to the three original tasks. Versions 1.0.0 and 1.2.2 were respectively available in the Android Google Play Store and iOS App Store at the time of writing and were used in this study by enrolled participants.

**Figure 2.**
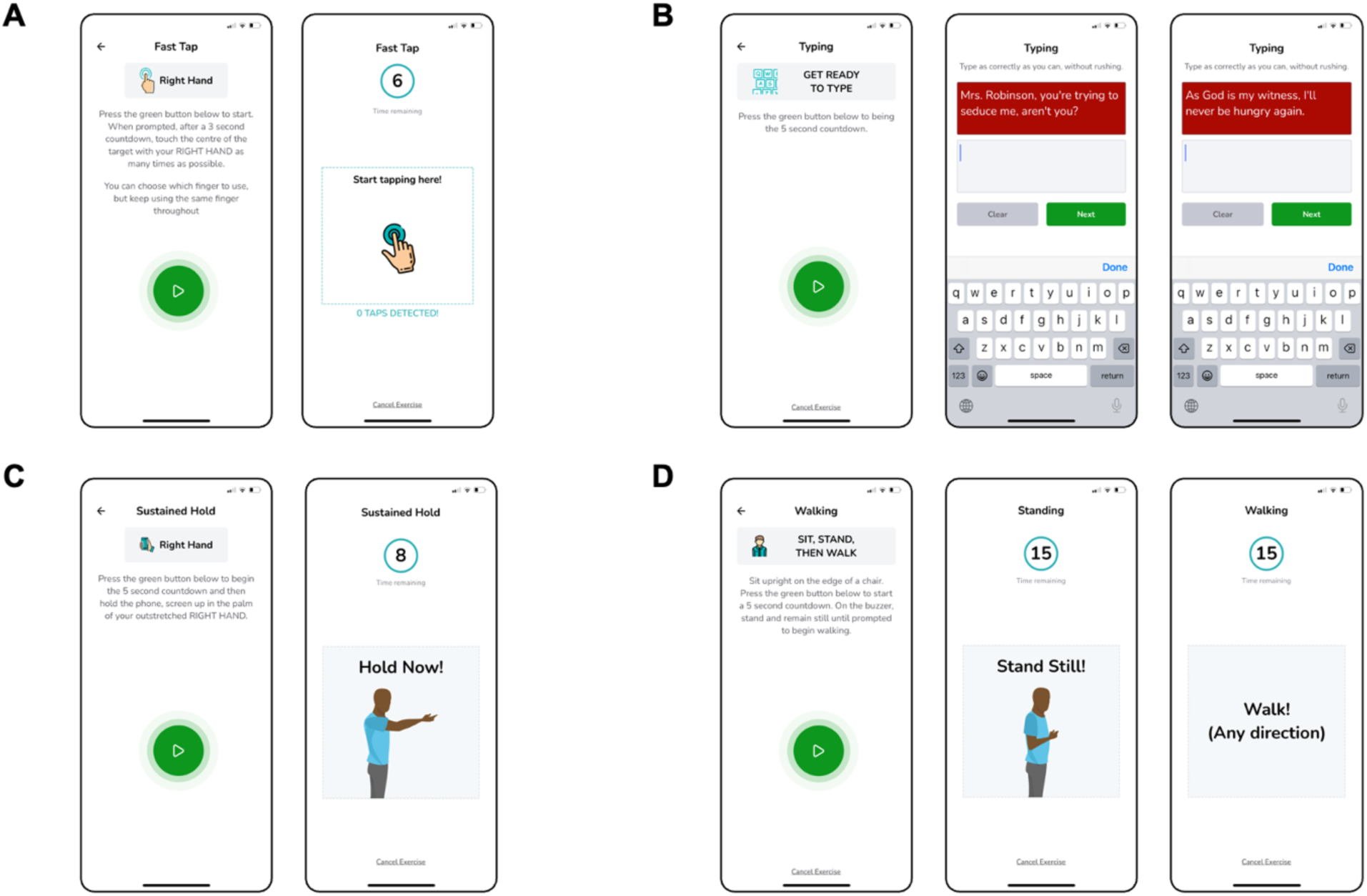
Schematic illustrations of the MoveMed battery of performance outcome measures. A: The six-second fast-tap test. B: The two-stage typing test. C: The eight-second sustain-hold test. D: The 15-second stand and walk test.

The Fast Tap Test is a PerfO task that assesses finger dexterity through an interactive, six-second smartphone task. Users are shown a demonstrative cartoon (Figure 2A) and instructed to ‘touch the centre of the target with [each] hand as many times as possible’. In-app video demonstration is also available. The construct (finger dexterity) is assessed by measuring the speed, accuracy, and efficiency of finger tapping as continuous variables and analysing them as a panel of measures. Content and construct validity were previously assessed by Yanez Touzet et al (5). In this study, tap count and latency were used as reflective measures of finger dexterity.

The Typing Test is another PerfO task that assesses finger dexterity through an interactive, two-stage smartphone task. Users are shown a demonstrative cartoon (Figure 2B) and instructed to ‘type as correctly as they can, without rushing’. In-app video demonstration is also available. The construct (finger dexterity) is assessed by measuring speed, accuracy, and efficiency of typing as continuous variables and analysing them as a panel of unidimensional measures. Content and construct validity were previously assessed in Yanez Touzet et al (5). In this study, typing speed was used as a reflective measure of finger dexterity.

The Hold Test is a PerfO task that assesses upper limb stability through an interactive, eight-second smartphone task. Users are shown a demonstrative cartoon (Figure 2C) and instructed to ‘hold the phone, screen up in the palm of [their] outstretched hand’. In-app video demonstration is also available. The construct (upper limb stability) is assessed by measuring the involuntariness, rhythmicity, and oscillation of the upper limbs as continuous variables and analysing them as a multidimensional Stability Score. Content and construct validity were previously assessed in Yanez Touzet et al (5). In this study, the Stability Score was used as a reflective measure of upper limb stability (5).

The Stand and Walk Test is a PerfO task that assesses gait through an interactive, two-stage smartphone task. During the first stage, users are instructed to ‘sit upright on the edge of a chair [and to] press the green button [when they are ready to] stand and remain still’. During the second stage, users are instructed to ‘walk [in] any direction’. In-app cartoon and video demonstration are also available (Figure 2D). The construct (gait) may then be assessed by measuring standing or walking as continuous variables and analysing them as multidimensional measures. Content and construct validity were previously assessed in Yanez Touzet et al (5). In this study, cadence was used as a reflective measure of gait.

### Patient-report outcomes

Two patient-reported outcome measures (PROM’s) were used as descriptors for DCM: the P-mJOA and the WHOQOL-Bref.

The P-mJOA score is a patient-reported questionnaire that assesses neuromuscular function in DCM across four items: motor function of the upper and lower extremities (MDUE and MDLE, respectively), sensory function of the upper extremities, and sphincter function (8). Responses are scored on an ordinal scale per item and presented as both a panel of scores and an unweighted sum-total score. The P-mJOA was selected due to the existence of systematic assessment of construct validity (*r* > 0.5) and feasibility in DCM (3) and due to the use of its clinically reported analogue as the current gold standard. The P-mJOA was favoured over the mJOA since it is intended to be a truly patient-reported equivalent of the mJOA, which can be understood by individuals with no medical knowledge or training (9).

The WHOQOL-Bref is a patient-reported questionnaire that assesses quality of life across 26 items grouped into four domains: physical health (PH), psychological health (PS), social relationships (SR), and environment health (EH) (10). Responses are scored on a five-point ordinal scale per item and presented as a panel of sum-total scores. Responses to two items may, furthermore, be presented individually to give insight into the respondent’s global perception of their quality of life and their quality of health. These were presented in writing to describe the population’s characteristics but were not considered robust enough to warrant correlation analysis. The WHOQOL-Bref was selected due to the existence of systematic assessments of validity, reliability, and responsiveness in traumatic brain injury (11), Parkinson’s disease (12), and DCM (3). It was also favoured over the 36-Item Short Form (SF-36) Health Survey due to its relative brevity and over the EuoQOL Five Dimensions (EQ-5D) Questionnaire due to licensing restrictions.

### Statistical analysis

The COSMIN manual defines reliability as ‘the degree to which scores for patients who have not changed are the same for repeated measurement under several conditions’ (13). Reliability may be formally assessed through test-retest reliability and measurement error.

In this study, we assessed reliability using test-retest reliability as the primary outcome and measurement error as a secondary outcome. Intraclass correlation coefficients of agreement (ICC_agreement_) were computed to assess the test-retest reliability of the PerfO’s continuous results. Standard error of agreement of the mean (SEM_agreement_) and the smallest detectable change of agreement (SDC_agreement_) were subsequently computed to assess measurement error of the PerfO’s continuous results. SDC_agreement_ was computed using the formula: 1.96 × √2 × SEM_agreement_ (14).

Data from the 12 weeks of the study were included due to the repeated-measures nature of the reliability analysis. Individuals were considered eligible for analysis if: (a) there were at least two test repeats per PerfO; (b) their P-mJOA scores at weeks 6 and 12 did not change clinically; and (c) there were no contextual anomalies in the time series for that individual, i.e., no subset of data points within the time series deviated significantly from the entire data set (15). Clinical change was assessed using the minimum clinically important difference (MCID) of the P-mJOA: namely, ‘between 1 and 2 points’ from baseline, as in Tetreault et al (16). Since the MCID in DCM is a range and varies with disease severity, reliability was studied and presented in two subgroups, MCID ≤1 and MCID 2, to aid with clinical interpretation.

All data were analysed using Python 3.10.12. ICC_agreement_ coefficients were calculated for successive repeats of the MoveMed PerfOM’s using the single random raters model from the `pingouin` package (17). Missing data were not imputed; thus, time periods with least missing data were selected for each PerfOM to mitigate the impact of missing data (17). SEM_agreement_ and SDC_agreement_ were calculated using the same series of successive repeats as the ICC_agreement_ coefficients.

### Risk of bias assessment

The COSMIN Risk of Bias checklist (13) was used to assess the methodological quality of test-retest reliability and measurement error.

### Overall assessment

Overall assessments of reliability were made using a panel of ratings. As in COSMIN (13), ICC_agreement_ were converted into ratings by comparing results to a critical threshold of 0.7. ICC_agreement_ ≥ 0.7 were rated ‘sufficient’. ICC_agreement_ < 0.7 were rated ‘insufficient’.

## RESULTS

27 participants with DCM enrolled in the prospective and decentralised EMPOWER study (Figure 3), principally via advertisement through Myelopathy.org, a DCM charity (6, 7). Twenty participants (74%, 20/27) were deemed ineligible for analysis due to the absence of a completed follow-up questionnaire (Figure 3). Seven (26%, 7/27) met all criteria for analysis (Table 1). From these, 7/7 (100%) were assigned to the MCID ≤2 group and 5/7 (86%) to the more conservative MCID ≤1 group.

**Figure 3.**
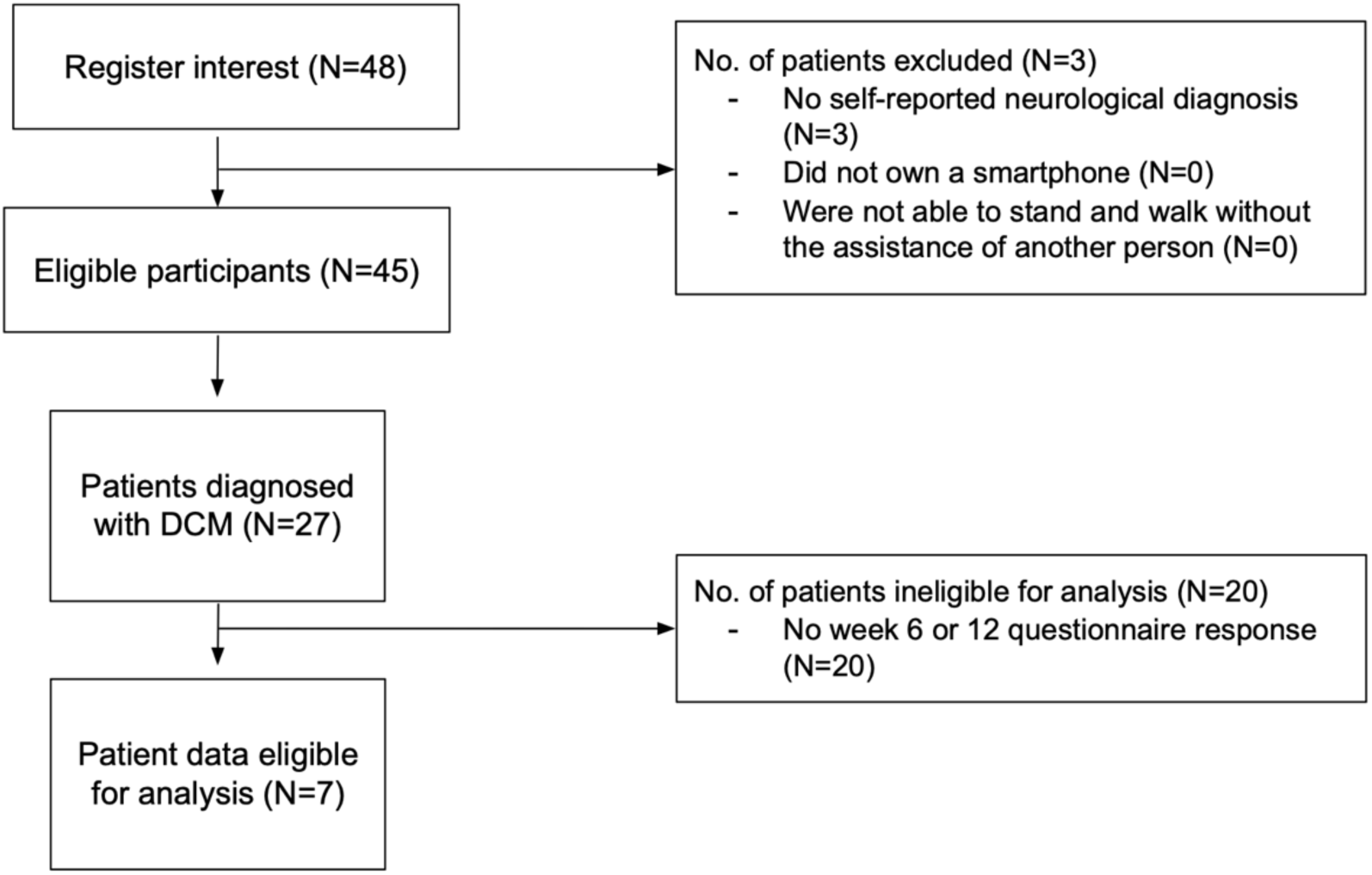
STROBE diagram.

**Table 1.** Characteristics of study participants.

| Feature | MCID≤1 | MCID≤2 |
| --- | --- | --- |
| No. of participants | 5 | 7 |
| Age | 62.0 (18.4) | 56.5 (12.4) |
| P-mJOA (reference range: 0–18) | 10.8 (2.3) | 12.5 (3.6) |
| MDUE (reference range: 0–5) | 3.4 (1.1) | 3.4 (1.1) |
| MDLE (reference range: 0–7) | 3.4 (0.9) | 4.1 (1.6) |
| SDUE (reference range: 0–3) | 2.0 (0.7) | 2.1 (0.8) |
| SD (reference range: 0–3) | 2.0 (0.0) | 2.4 (0.5) |
| WHOQOL-Bref | — | — |
| Overall QOL (reference range: 1–5) | 2.6 (0.9) | 3.1 (1.0) |
| Overall health (reference range: 1–5) | 2.6 (0.5) | 3.0 (0.8) |
| EH (reference range: 8–40) | 26.2 (5.2) | 27.1 (4.8) |
| PH (reference range: 7–35) | 17.8 (6.6) | 19.9 (6.3) |
| PS (reference range: 6–30) | 18.6 (3.4) | 20.1 (3.5) |
| SR (reference range: 3–15) | 9.8 (1.8) | 9.4 (1.6) |
| MoveMed Fast Tap Test Count | — | — |
| Dominant hand | 24.7 (5.9) | 24.7 (5.9) |
| Non-dominant hand | 25.7 (8.3) | 24.0 (7.9) |
| MoveMed Fast Tap Test Inter-Tap Duration (s) | — | — |
| Dominant hand | 0.19 (0.12) | 0.18 (0.10) |
| Non-dominant hand | 0.15 (0.04) | 0.15 (0.05) |
| MoveMed Hold Test Stability Score (%) | — | — |
| Dominant hand | 83.3 (9.9) | 72.1 (23.1) |
| Non-dominant hand | 80.5 (15.7) | 70.1 (25.7) |
| MoveMed Typing Test Speed (keys per second) | 1.60 (0.55) | 1.55 (0.47) |
| MoveMed Stand and Walk Test Cadence (steps per min) | 59.6 (31.7) | 64.6 (30.1) |

Data reported as *n* or mean (standard deviation) unless otherwise specified. Ranges reported in manuscript body.
EH: Environmental health, MDLE: Motor dysfunction of the lower extremity, MDUE: Motor dysfunction of the upper extremity, P-mJOA: Patient-derived modified Japanese Orthopaedic Association, PH: Physical health, PS: Psychological health, QOL: Quality of life, SD: Sphincter dysfunction, SDUE: Sensory dysfunction of the upper extremity, SR: Social relationships, WHOQOL-Bref: World Health Organization QOL Brief Version

In both groups, participants averaged 60 years old (MCID ≤2: 56.6, SD 12.4; MCID ≤1: 62.0, SD 18.4; Table 1). DCM severity ranged from mild to severe in MCID ≤2 (P-mJOA total score: 8–18) and from moderate to severe in MCID ≤1 (P-mJOA total score: 8–14). Upper limb motor function ranged from none to ‘unable to eat with spoon but able to move hands’ in both groups (P-mJOA MDUE subscore: 2–5). Lower limb motor function ranged from none to ‘able to move legs but unable to walk’ in MCID ≤2 (P-mJOA MDLE subscore: 2–7) and from ‘able to walk up-&/or downstairs w/aid of a handrail’ to ‘able to move legs but unable to walk’ in MCID ≤1 (P-mJOA MDLE subscore: 2–4). In both groups, overall QOL perception ranged from ‘good’ to ‘poor’ (WHOQOL Overall QOL: 2–4), whilst overall health perception ranged from ‘satisfied’ to ‘dissatisfied’ in MCID ≤2 (WHOQOL Overall health: 2–4) and from ‘satisfied’ to ‘neither poor nor good’ in MCID ≤1 (WHOQOL Overall health: 2–3).

On MoveMed, participants performed similarly across groups (Table 1). In the Fast Tap Test, participants tapped the screen between 24 to 25 times and waited around 1500–1900 ms between taps on average. In the Typing Test, they typed around 1.5 keys per second. In the Hold Test, arm stability averaged 70% to 80% points and in the Stand and Walk Test, cadence averaged 60 steps per minute.

### Reliability

ICC_agreement_ coefficients for both groups are reported in Table 2. Coefficients for the Hold and Typing Tests were ≥0.7 in both groups. Coefficients for the Fast Tap Test Count were also ≥0.7 in both groups. Coefficients for the Fast Tap Test Inter-Tap Duration, however, were ≥0.7 in the MCID ≤1 group only. Coefficients for the Stand and Walk Test were 0.68 in MCID ≤1 and 0.62 in MCID ≤2. Higher coefficients were generally seen in the more conservative test-retest stability group (i.e., MCID ≤1).

**Table 2.** ICC_agreement_ coefficients for the MoveMed outcome measures.

| Outcome measure | Endpoint | Result | N | Rating* | ROB |
| --- | --- | --- | --- | --- | --- |
| **MCID ≤1** |  |  |  |  |  |
| Fast Tap Test | Count | DH: 0.94, NH: 0.92 | 5 | DH: +, NH: + | No |
|  | Inter-Tap Duration | DH: 0.97, NH: 0.84 | 5 | DH: +, NH: + | No |
| Hold Test | Stability Score | DH: 0.89, NH: 0.95 | 5 | DH: +, NH: + | No |
| Typing Test | Speed | 0.95 | 4 | + | No |
| Stand and Walk Test | Cadence | 0.68 | 4 | – | No |
| **MCID ≤2** |  |  |  |  |  |
| Fast Tap Test | Count | DH: 0.88, NH: 0.91 | 7 | DH: +, NH: + | No |
|  | Inter-Tap Duration | DH: 0.61, NH: 0.68 | 7 | DH: –, NH: – | No |
| Hold Test | Stability Score | DH: 0.77, NH: 0.75 | 7 | DH: +, NH: + | No |
| Typing Test | Speed | 0.98 | 6 | + | No |
| Stand and Walk Test | Cadence | 0.62 | 5 | – | No |

\* '+' = Sufficient, '–' = Insufficient
DH: Dominant hand, NH: Non-dominant hand, ROB: Risk of bias

### Measurement error

SEM_agreement_ and SDC_agreement_ for both groups are reported in Table 3. In MCID ≤1, SEM_agreement_ values for the Fast Tap, Hold, Typing, and Stand and Walk Tests were, respectively: ±1 tap, ±1–3% points, ±0.06 keys per second, and ±10 steps per minute. In MCID ≤2, SEM_agreement_ values were: ±1 tap, ±2–4% stability score points, ±0.06 keys per second, and ±10 steps per minute. SDC_agreement_ values (i.e., thresholds above or below which change scores can be considered real change and not change due to measurement error (13)) were: ±3 taps, ±4–7% points, ±0.2 keys per second, and ±27 steps per minute in MCID ≤1, and ±3–7 taps, ±7–10% points, ±0.2 keys per second, and ±27 steps per minute in MCID ≤2. Lower measurement errors were generally seen in the more conservative MCID ≤1 group for the Fast Tap and Hold Tests.

**Table 3.**
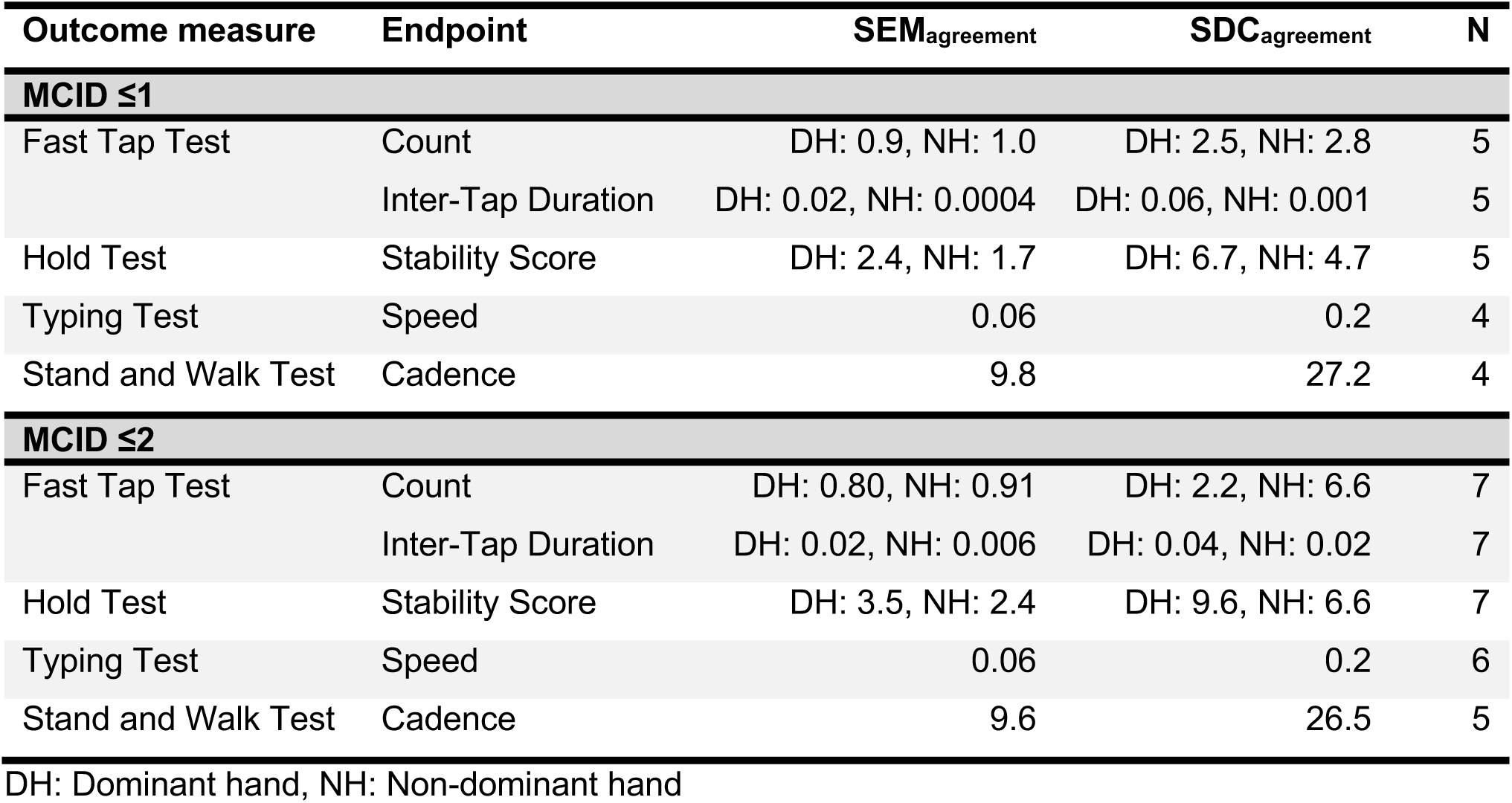
Measurement error metrics for the MoveMed outcome measures.

| Outcome measure | Endpoint | SEM<sub>agreement</sub> | SDC<sub>agreement</sub> | N |
| --- | --- | --- | --- | --- |
| **MCID ≤1** |  |  |  |  |
| Fast Tap Test | Count | DH: 0.9, NH: 1.0 | DH: 2.5, NH: 2.8 | 5 |
|  | Inter-Tap Duration | DH: 0.02, NH: 0.0004 | DH: 0.06, NH: 0.001 | 5 |
| Hold Test | Stability Score | DH: 2.4, NH: 1.7 | DH: 6.7, NH: 4.7 | 5 |
| Typing Test | Speed | 0.06 | 0.2 | 4 |
| Stand and Walk Test | Cadence | 9.8 | 27.2 | 4 |
| **MCID ≤2** |  |  |  |  |
| Fast Tap Test | Count | DH: 0.80, NH: 0.91 | DH: 2.2, NH: 6.6 | 7 |
|  | Inter-Tap Duration | DH: 0.02, NH: 0.006 | DH: 0.04, NH: 0.02 | 7 |
| Hold Test | Stability Score | DH: 3.5, NH: 2.4 | DH: 9.6, NH: 6.6 | 7 |
| Typing Test | Speed | 0.06 | 0.2 | 6 |
| Stand and Walk Test | Cadence | 9.6 | 26.5 | 5 |

DH: Dominant hand, NH: Non-dominant hand

### Risk of bias assessment

No risk of bias factors from the COSMIN Risk of Bias checklist were recorded, which was equivalent to a ‘very good’ quality rating for methodological quality (13).

### Overall assessment

Reliability ratings for both groups are also reported in Table 2. ICC_agreement_ coefficients were converted into ratings by comparing results to a critical threshold of 0.7. All unidimensional tests obtained sufficient ratings for test-retest reliability across groups. In MCID ≤1, sufficient ratings were obtained for the performance of the Fast Tap, Hold, and Typing Tests. In MCID ≤2, sufficient ratings were obtained for the performance of the Hold and Typing Tests, and the total count of the Fast Tap Test. The performance of the multidimensional Stand and Walk Test was the only test that did not achieve a sufficient rating, due to ICC_agreement_ coefficients of 0.68 in MCID ≤1 and 0.62 in MCID≤2. Taken together, these data provide ‘very good’ quality evidence for the overall reliability of the PerfOM’s in the assessment of DCM.

## DISCUSSION

The primary purpose of this study was to assess the reliability of MoveMed, a battery of PerfOM’s in a mobile app. Two aspects were evaluated: test-retest reliability and measurement error. All PerfOM’s demonstrated moderate-to-excellent test-retest reliability coefficients (Table 2) and low measurement errors (Table 3). These findings are important as they confirm that MoveMed’s PerfO’s remain stable when DCM remains stable, and provide reference values to discern real vs. random change in each PerfO (13).

Software applications are increasingly being used to administer PerfOM’s in medicine (18). However, most digital development to date has focused on running PerfOM’s on wearable technology over smartphones. Smartwatches, for instance, have been frequently used to measure activity and movement, but test-retest reliability has been found to be insufficient at worst, and variable at best (19, 20). Smartphone testing has recently emerged as a more reliable alternative (21, 22). In our experience, this is because smartphone testing (a) focuses on narrower constructs, (b) standardises task performance (i.e. conducts task in a controlled or stereotyped environment), and (c) controls for environmental variables better as a result. These principles incidentally reflect John Macleod’s fundamentals of good clinical examination and have been intentionally designed into MoveMed to ‘improve the reliability and precision of… clinical assessment’ (23).

Few PerfOM’s are investigated for reliability thoroughly, including smartphone applications (13, 24-28). Test-retest coefficients are often computed but measurement errors are often ignored. This is detrimental to clinical practice as it leaves clinicians unable to confidently differentiate real vs. random changes in outcomes. Furthermore, when reported in isolation, test-retest coefficients run the risk of optimistic interpretation. The basis for this is the truism in Portney (29) that ‘reliability cannot be interpret as an all-or-none condition [but rather as] attained to varying degrees’. This leads to graded interpretation of coefficients, on an (admittedly) arbitrary scale of ‘poor’ (<0.50) to ‘moderate’ (0.50–0.75) to ‘good’ (>0.75) (29) and even ‘excellent’ (>0.90) (30). These criteria, however, do not help clinicians judge whether tools are ‘sufficiently’ reliable for use or not. Consensus guidance has been developed for this purpose and recommend using an all-or-none cut-off of 0.70 (13, 31). In our study, all PerfOM’s were deemed sufficiently reliable using this cut-off, bar the multidimensional Stand and Walk Test, where the ICC_agreement_ coefficient was 0.68 in the conservative [MCID <1] group. We attribute this deficit primarily to the multidimensional nature of the construct and optimisation is underway to improve control of random error through greater standardisation in an updated version of MoveMed, due to be released shortly.

This study computed measurement errors, which enable researchers to interpret readings (13). Our findings suggest that changes above or below these thresholds can be considered real change in individual user performance, and not change due to measurement error: (a) changes above or below seven taps in the Fast Tap Test; (b) changes above or below 10 stability score points in the Hold Test; (c) changes above or below 0.2 keys per seconds in the Typing Test; and (d) changes above or below 27 steps per minute in the Stand and Walk Test (Table 3). A further study will be needed to elucidate the clinical significance of these thresholds (i.e., what is the magnitude of change which should trigger a change in clinical care and/or is considered meaningful to the patient). This is referred to as the MCID and may be different to the SDC.

An important strength of this study is its design by individuals with formal training in clinimetrics. This is reflected in the absence of risk of bias factors from the COSMIN checklist in Table 2, and the study’s reporting. Use of the COSMIN manual is strongly encouraged by the authors, as its standards are considered to be a cornerstone in clinimetric validation and overlap importantly with industry guidance from the United States Food and Drug Administration (32, 33). Another strength in this study was the use of PerfOMs that can collect several test-retest repeats quickly, ecologically, and longitudinally. This means that constructs should be captured more precisely, more reflective of pathology in the patient’s natural environment, and potentially more responsive to intervention.

Despite its conscientious design, this study has limitations. First, several participants were excluded from analysis. Three quarters of the sample (74%, 20/27) were excluded due to the absence of follow-up questionnaire responses and up to a tenth (11%, 3/27) due to missing repeated-measures data (17). We attribute the former exclusion to the burden of repeating PROM completion for patients, and the latter to heterogenous task adherence. To maximise the clinical relevance of our findings, we stratified the included sample into demonstrably stable groups: namely, a conservative MCID ≤1 group and a less conservative MCID ≤2 group. This combination was chosen as the MCID of the mJOA is estimated between 1 and 2; recognised to vary depending on the severity of the individual’s disease and the method by which it is estimated (16). That the reliability was increased for the MCID ≤1 group compared to MCID ≤2 group, further supports the reliability of the measure itself. Whilst MoveMed data was longitudinally available for 100% (27/27) of the sample, clinical stability could only be demonstrated in a subset and so it was deemed safer and more beneficial to report on the subset. In the next version of the app, in-app reminders and gamification will improve homogeneity of adherence and follow-up.

Second, standards for patient-reported methods were adapted to assess performance-based methods. This was done to overcome the absence of standardised criteria in this field and because there is precedent for it in Terwee et al (24) and the COSMIN manual (13). Third, people with a severe form of the disease may have been excluded from enrolment. This would be due to the exclusion of individuals that were unable to stand and walk without the assistance of another person. The potential risks of remote participation in this subset of individuals, however, were deemed to outweigh the benefits by the ethical committee. Further in-person research could address this limitation in the future.

## CONCLUSIONS

The current study provides initial evidence for the reliability of the MoveMed PerfOM’s in the context of adults living with DCM in community.

## DATA AVAILABILITY STATEMENT

All data relevant to the study are included in the article.

## CONTRIBUTORS

BMD and AYT conceived the study and are the guarantors. AYT, BMD, TH, and ZR collected and analysed the data. AYT and BMD wrote the manuscript. AK, SY, DBA, IL, ML, GG, and MRNK provided critical and independent appraisal of the methods, data, and manuscript. All authors reviewed and critically revised the manuscript prior to submission.

## Data Availability

All data produced in the present work are contained in the manuscript.

## ACKNOWLEDGEMENTS

We thank the ongoing support of all participants and stakeholders, including Myelopathy.org (DCM Charity; www.myelopathy.org).

